# Targeting the gut microbiome in allogeneic hematopoietic stem cell transplantation

**DOI:** 10.1101/2020.04.08.20058198

**Authors:** Marcel A. de Leeuw, Manuel X. Duval

**Affiliations:** GeneCreek

## Abstract

**Background:** Gut microbiota (GM) composition has been associated with acute Graft-versus-Host Disease (aGvHD), however, current knowledge is insufficient to target the GM.

**Methods:** A relevant series of microbiome data sets were combined and reanalyzed, with resolution of species level changes in the GM.

**Results:** GM composition was found strongly correlated with aGvHD status after one month post stem cell infusion (R2=0.51). The predicted average biological safety level, indicative of antibiotic resistance, was found increased in aGvHD cases (p=1.6E-4). Using in silico modelling, we formulated a probiotic composition putatively competing with mortality and aGvHD associated species.

**Implications:** Supplementation with *Bifidobacterium longum* and *Bifidobacterium breve* is proposed as a prophylactic treatment alongside with prebiotics and an adapted antibiotics course.

## Introduction

Allogeneic Hematopoietic Stem Cell Transplantation (allo-HSCT) is a curative option for many patients with high-risk hematopoietic malignancies and hematological disorders. The success of allo-HSCT can be hampered by a process in which donor-derived T cells recognize host healthy tissue as non-self, causing an immunemediated complication known as acute Graft-versus-Host Disease (aGvHD), predictive for morbidity and mortality of the patients. Even among transplants sourced from HLA-matched siblings, aGvHD occurs in 40% of recipients and reaches 50-70% with unrelated donor HSCT, yet less than half of the patients who develop aGVHD experience a response [6]. Bacterial LPS has been proposed as a triggering factor for aGvHD, based on murine models [7]

In patients undergoing allo-HSCT, antibiotics are routinely prescribed to reduce the risk of opportunistic infections. Antibiotics can profoundly impact gut microbiome (GM) composition and it has been shown broad spectrum antibiotics used after allo-HSCT increase the risk of aGvHD in mice and humans [8, 9]. On the other hand, gut decontamination (GD), i.e. the complete suppression of the GM, has been shown to prevent aGvHD following allo-HSCT, but is difficult to achieve [10, 11]. As a matter of fact, no standardized protocol for prophylactic and peri-transplant antibiotic treatment has been established as standard of care across transplantation centers.

It has been reported that autologous fecal microbiota transfer (auto-FMT) can in some instances restore the baseline GM after antibiotics courses applied during the allo-HSCT [1, 12]. Of note, this baseline GM is potentially already disturbed, as a possible side effect of chemotherapy [13]. Hence some centers practice probiotics and 3^rd^ party FMT reconditioning of GM once the antibiotic treatment has been ended [11, 14]. Third party FMT is also practiced with success for the treatment of steroid refractory aGvHD [15–18].

A lower GM diversity index at the time of engraftment (neutrophil recovery) has been associated in multiple studies with increased incidence of intestinal aGvHD disease and transplant related mortality (TRM) [19–24]. *Enterococci* have been proposed as a landmark for aGvHD whether induced by antibiotics or not [25, 26], whereas intestinal *Blautia* has been found associated with reduced death from aGvHD [27]. It has also been reported that at the time of engraftment, decreased *Lachnospiraceae* and *Ruminococcaceae* and increased *Enterobacteriaceae* correlate with aGvHD development and a lowered Treg/Th17 ratio [22].

We combined a series of relevant microbiome studies, available in the form of raw 16S data, in order to increase statistical power and investigate detailed baseline and aGvHD onset microbiome composition in relation to therapeutic outcome. At baseline we found that pre-conditioning regime is associated with the GM composition and that composition is correlated with survival. We found that from one month post stem cell infusion onward, aGvHD cases and non-aGvHD controls have distinguishable GM. We predicted relative gene abundance differences for several metabolites of interest. Finally, we predicted qualified presumption of safety (QPS) species which could potentially be used as prophylaxis to control detrimental species, respect control-associated species and improve outcomes.

## Materials & Methods

The materials summarized in Table 1 have been made available in the form of raw 16S data sets in the short read archive (SRA). Five of the six collected data sets have been made available as part of scientific publication. Four of the data sets are from longitudinal GM studies. Only one data set, ERP017899, was published with extensive per sample metadata, but for all but one data set, SRP162022, aGvHD status information was available.

**Table 1:** Prospective data sets used in the study. SRA: short read archive id, Qiita: Qiita index number, https://qiita.microbio.me, N: number of patients, n: number of samples, 16S: variable regions covered. *unpublished.

| BioProject | SRA | Qiita | N | n | 16S | publ. | study details |
| --- | --- | --- | --- | --- | --- | --- | --- |
| PRJNA491657 | SRP162022 |  | 736 | 2,794 | V4-V5 | [1] | 735 HCT patients with auto-FMT, longitudinal |
| PRJEB16057 | ERP017899 | 10,564 | 48 | 48 | V4 | [2] | 25 GvHD patients, 23 matched controls and 16 donors, baseline |
| PRJNA602484 | SRP243841 |  | 61 | 61 | V3-V4 | * | 31 GvHD patients and 30 matched controls, aGvHD onset |
| PRJEB23820 | ERP105598 |  | 26 | 84 | V3-V4 | [3] | 15 GvHD patients and 11 matched controls, longitudinal |
| PRJNA592853 | SRP234378 |  | 20 | 104 | V3-V4 | [4] | 10 GvHD patients and 10 matched controls, longitudinal |
| PRJEB28845 | ERP111102 |  | 19 | 246 | V4 | [5] | 4 GvHD patients and 15 matched controls, longitudinal |

### Data analysis

Amplicon Sequence Variants (ASVs) were generated with the R Bioconductor package dada2, version 1.12.1 with recommended parameters [28], involving quality trimming, discarding of sequences with N’s, assembly of forward and reverse sequences and chimera removal, as described previously [29]. Further analysis involved multiple alignment with mafft, version 6 [30] and approximately-maximum-likelihood phylogenetic tree generation with FastTreeMP, version 2 [31].

Taxonomic classification of ASVs was performed by an in-house Python and R program using random forest based supervised learning on the Ribosomal Database Project (RDP) release 11.5. Phenotypic content of microbiomes was inferred using species-level taxonomic assignments only and an in-house phenotypic database which has been initiated with information from Bac-Dive [32] and the IJSEM journal [33] and has been hand-curated since. Resulting classifications and microbiome phenotypes are available from the Github repository https://github.com/GeneCreek/GvHD-manuscript in the form of Phyloseq R data objects.

Random forest survival analysis was carried out with the R package ranger, version 0.12.2.

aGvDH status regression analysis, with relative abundance of taxa resolved at the species level as independent variables, was performed using the R package relaimpo [Groemping, Ulrike [34].

Relative gene abundance prediction was performed using in-house R code, correcting for 16S copy numbers using the rrnDB version 5.4 [35] and using UniProt [36] provided per species non-redundant proteome counts and per species gene counts.

Co-exclusion and co-occurrence between species for probiotics composition were computed using χ_2_ testing on detectable presence of species in samples (n=21,197) from a set of clinical- and crowd sourced 16S studies, all performed on the Illumina platform, Table 1 and supplemental Table S2.

Downstream analysis scripts covering the figures presented in the manuscript are available from https://github.com/GeneCreek/GvHD-manuscript.

## Results

### Overall GM composition evolution across allo-HSCT

Data set SRP162022 comprises patients undergoing allo-HSCT (n=736) of which 14 received an auto-FMT 49 days after stem cell infusion, Fig. 1. GvHD status of patients was not available in the metadata for this data set. Recovery of Shannon species diversity seems to be boosted through auto-FMT, whereas strict anaerobes were not recovered. The antimicrobial resistance of the microbiome decreased, as reflected by the average biological safety level (BSL), which means that a less pathogenic GM was recovered through auto-FMT.

**Figure 1:**
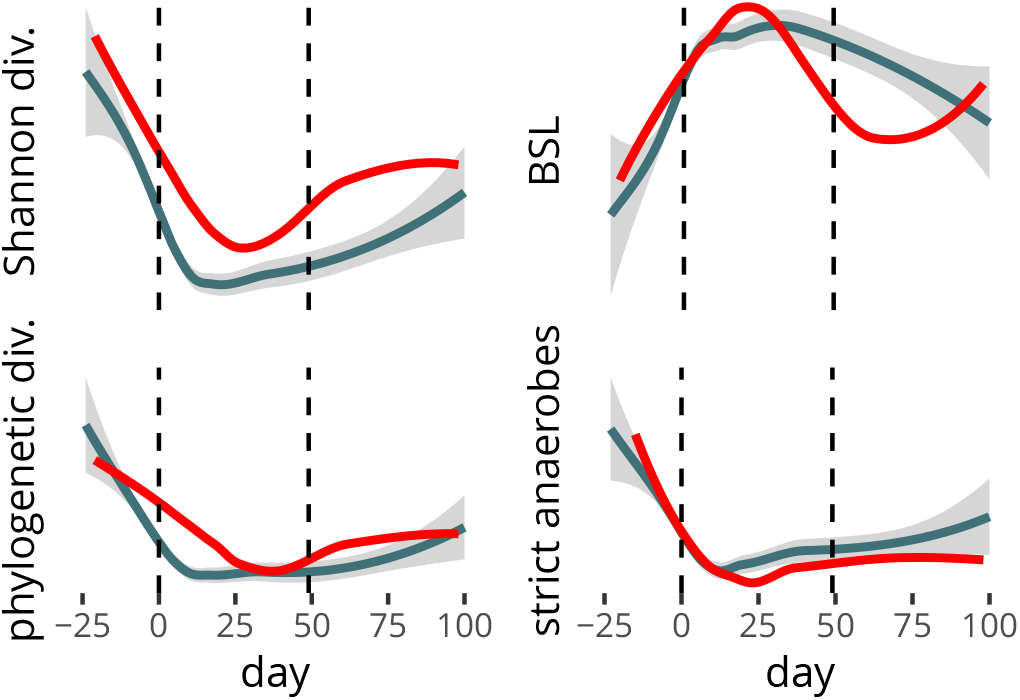
GM composition evolution across allo-HSCT. Data set SRP162022, 736 patients receiving stem cell infusion day 0. 14 patients (red) received an auto-FMT day 49. BSL: biological safety level.

### Correlation between conditioning and the GM

Data set ERP017899 contains immunosuppressive conditioning intensity specification and baseline GM samples of 48 patients before stem cell infusion. The conditioning was qualified as low, intermediate or high with few cases of high conditioning. We regrouped the intermediate level with the high level and tested various GM composition covariates comparing the two regimes. The Shannon species richness and the strict anaerobe proportion - the proportion of anaerobes that are obligate anaerobes - reached significance, Fig. 2. Both criteria also reach significance when comparing the baseline patient GM with donor GM, supplemental Fig. S1. Furthermore, the use of the immunosuppressant cyclosporine, which in data set ERP017899 is mutually exclusive with the use of tacrolimus, is correlated with the over-representation of gram-positive bacteria, supplemental Fig. S2.

**Figure 2:**
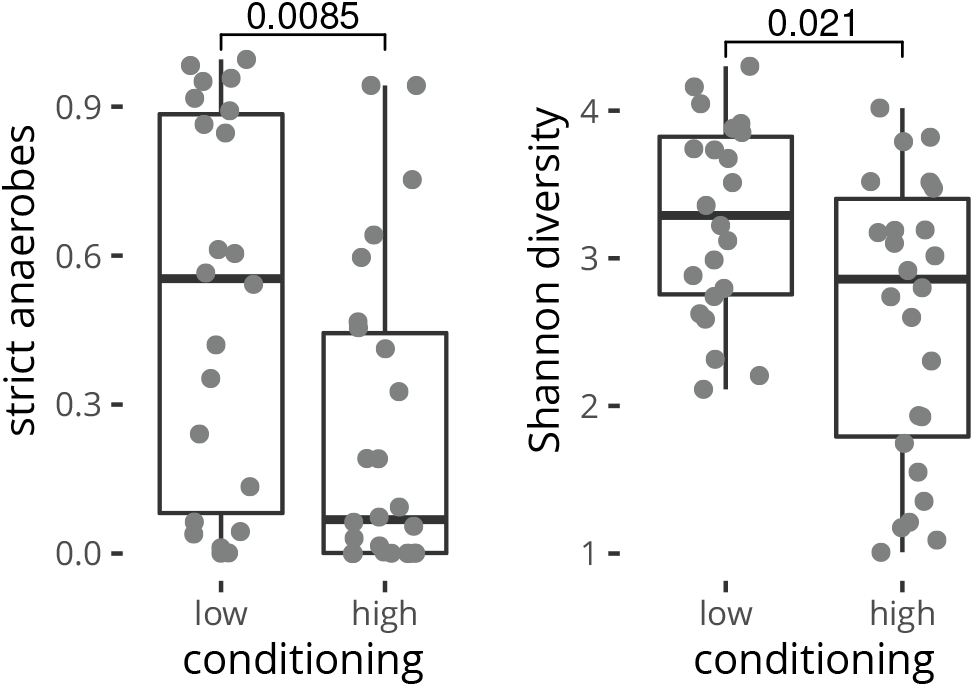
Baseline GM composition and conditioning level. Data set ERP017899, baseline samples of 41 patients who underwent allo-HSCT. Numbers reflect Wilcoxon signed rank test p-values.

### Baseline GM composition and survival

Data set ERP017899 contains baseline GMs and survival information for up to two years of 41 patients who underwent allo-HSCT. In brief, we fitted random survival forests (RSF) using log scaled relative abundances of taxa resolved at the species level as independent variables. Variable importance and associated p-values were estimated using permutations. We retained p<0.05 taxa as selected variables and the experience was repeated 50 times. Final models were built using 20 fold cross validation and a subset of taxa selected at least 10 times. Model performance was assessed using the integrated Brier score (IBS) and the best performing model was retained.

This model used 10 taxa, Fig. 3 and had an IBS of 0.153. All selected taxa had higher average relative abundance at baseline in patients who subsequently deceased during the two year censoring time span.

**Figure 3:**
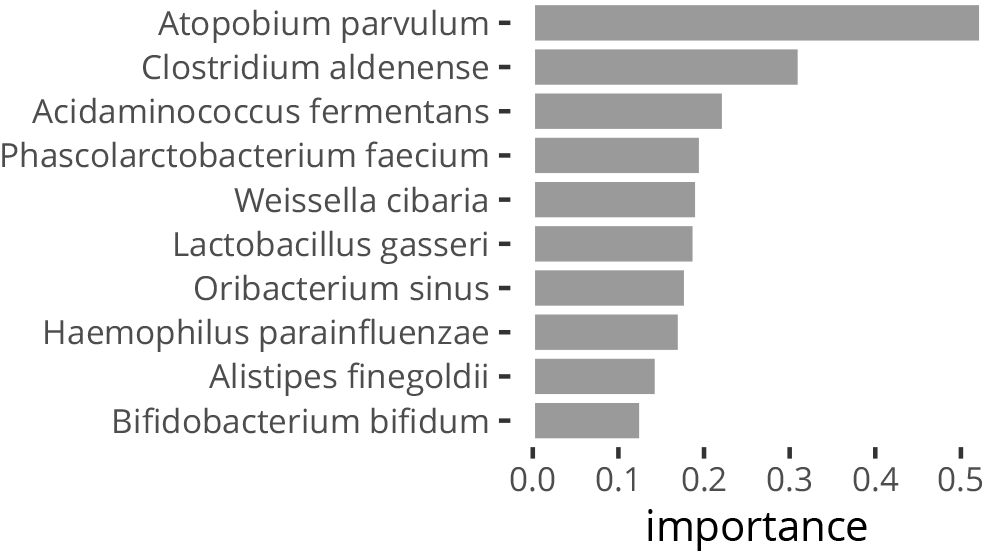
Top 10 variable importances estimated by the random survival forest models. Data set ERP017899, 41 patients who underwent allo-HSCT, with right-censoring of survival up to 2 years. All selected species are associated with mortality.

### aGvHD cases, controls and GM composition

We combined data sets SRP243841, ERP105598, SRP234378 and ERP111102 into a 84 patients and 178 samples data set comprising samples taken one month past stem cell infusion or beyond with matched controls. We defaulted to a case/control analysis not disposing of detailed aGvHD censoring information for these data sets. There is a significant difference in the inferred average biological safety level between the two patient groups, Fig. 4.

**Figure 4:**
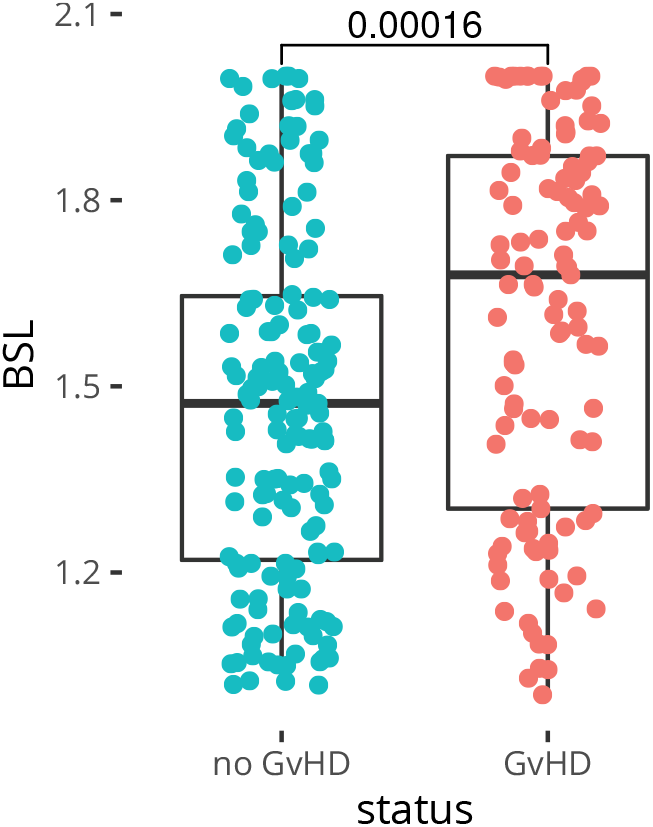
Biological safety level and aGvHD at onset. Combined data sets SRP243841, ERP105598, SRP234378 and ERP111102 - 84 patients and 178 samples. BSL: inferred average biological safety level of the microbiome. Numbers reflect Wilcoxon signed rank test p-values.

We performed the same analysis for donor GM samples but although we seem to see an identical trend, the low number of samples available (n=15) did not allow for the test to reach statistical significance, Fig. S3. Compositional differences between aGvHD cases and controls could also be documented at the species level. Using linear regression and relative importance analysis, we found that 20 taxa accounted for 51.3% (adjusted R^2^) of the aGvHG/non-aGvHD variability, Fig. 5. The linear model performs on the data set with a receiver operating characteristic (ROC) area under the curve (AUC) of 0.945, supplemental Fig. S4.

**Figure 5:**
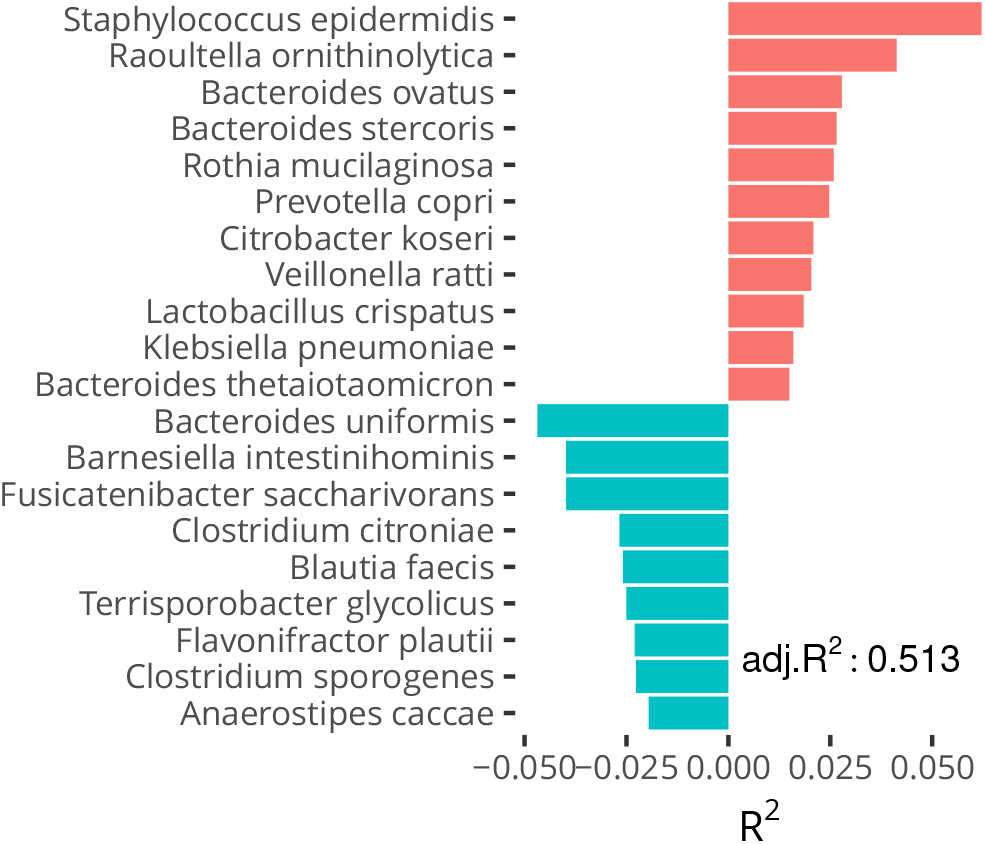
Relative importance of regressors explaining the aGvHD status. Combined data sets SRP243841, ERP105598, SRP234378 and ERP111102 - 84 patients and 178 samples.

### Immuno-modulating metabolites

Tryptophan derived aryl hydrocarbon receptor (AhR) ligands are critical for the maintenance of the intestinal epithelial barrier, and regulate innate and adaptive immunity [37, 38]. We found the predicted relative abundance of AhR ligand tryptamine producing aromatic acid decarboxylase (padI, ec:4.1.1.28) to be decreased in GvHD, Table S1.

Polyamines, i.e. putrescine, spermidine and spermine, enforce the epithelial barrier and modulate both innate and adaptive immunity [37]. We found two putrescine producing genes, biodegradative arginine decarboxylase (adiA, ec:4.1.1.19) and agmatine ureohydrolase (speB, ec:3.5.3.11) to be predicted decreased in aGvHD as compared to non-aGvHD controls, Table S1.

### In silico screening of the allo-HSCT GM

To investigate if qualified presumption of safety (QPS) species could be of therapeutic interest, we computed pairwise χ^2^ tests for all QPS species with mortality, aGvHD case and control associated species reported above, Fig. 6. We used 21,197 samples from several tens of clinical and crowd-sourced studies as the basis for the test, TAbles 1 and S2. Supplemental Fig. S5 provides the same analysis for control associated species and Fig. S6 provides pairwise χ^2^ tests results between QPS species, the former to assess respect of control associated species and the latter to assess compatibility of QPS species for combination therapy.

The top 4 species in Fig. 6 have an unfavorable profile with regards to control associated species, Fig. S5. However, *Bifidobacterium longum* and *Bifidobacterium breve* both are in exclusion with a number of case-associated species, are complementing each other in that respect and are found in mutual inclusion (Fig. S6), hence we predict the species are compatible and can be used in a combination therapy. *Bacillus clausii* is found in a position similar to *B. longum*. All three species are relatively respectful non-aGvHD control associated species, Fig S5, with three or fewer co-exclusions and substantial predicted synergy with control associated species for *B. longum*.

**Figure 6:**
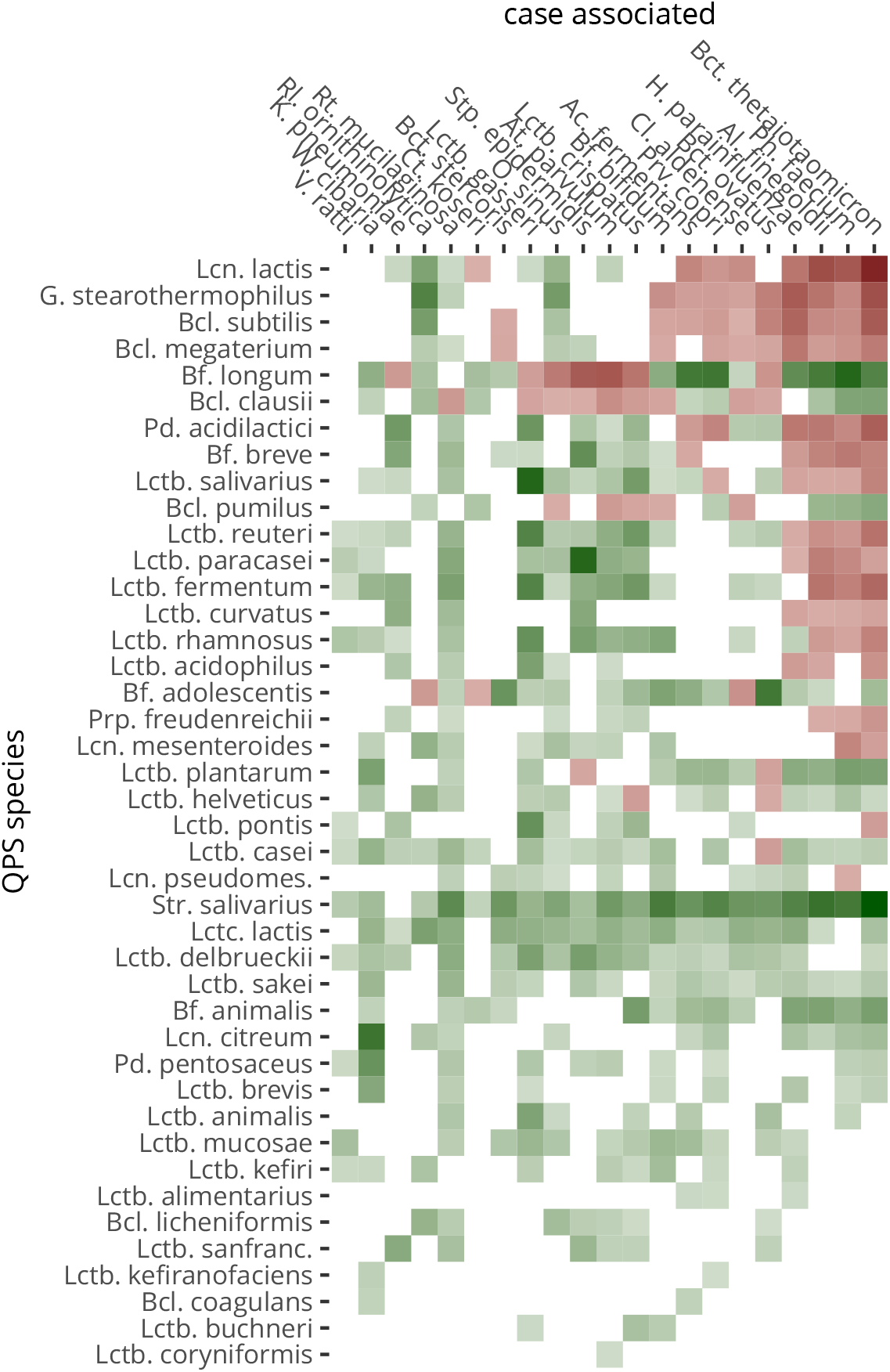
Co-exclusion by and co-occurrence with QPS species. Putative inhibition is in shades of red, potential synergy in shades of green. White reflect neutrality or too little combined prevalence to make a call. Genera are abbreviated as follows: **Bcl**.: *Bacillus*, **Bf.:** *Bifidobacterium*, **Gb.:** *Geobacillus*, **Lcn.:** *Leuconostic*, **Lctb.:** *Lactobacillus*, **Lctc.:** *Lactococcus*, **Pd.:** *Pediococcus*, **S.:** *Streptococcus*.

## Discussion

### Allo-HSCT conditioning and the GM

Our analysis of overall GM evolution across allo-HSCT shows that at baseline, i.e. after immunosuppressive conditioning, GM composition is already significantly altered, with notably decrease of strict anaerobes among which most butyrate producers are found. Cyclosporine seems to boost gram positive species, which could be beneficial in the scope of allo-HSCT, whereas its alternative tacrolimus does not. Furthermore, the latter is now known to be catabolized by intestinal Clostridia into less effective metabolites [41].

### Auto-FMT and allo-HSCT

If auto-FMT is to be practiced, the question arises when to source the samples. Sourcing after the initiation of immunosuppressive conditioning would avoid restoring butyrate producing species, which could be the safest option. GM production of butyrate benefits intestinal barrier function and decreases GvHD in a mouse model [42]. Notwithstanding this, it has been shown once aGvHD of the gut has occurred, butyrogens may impair recovery and lead to chronic GvHD [43]. Auto-FMT from our results not only restores a certain level of species diversity but also lowers the relative abundance of pathobionts as indicated by the predicted average biological safety level, which is an important feature not previously reported.

### PAMPs and aGvHD

Pathogen associated molecular patterns (PAMPs) function as potent stimulators for host and donor-derived antigen presenting cells (APCs) that in turn activate and amplify the responses of alloreactive donor T cells [44]. We predicted higher levels of antibiotic resistance of the GM to be associated with aGvHD cases. Increased relative abundance of resistance is associated with pathogens such as *Klebsiella pneumoniae* and *Staphylococcus epidermidis*. In addition, the efflux pumps responsible for antibiotic resistance are also capable of exporting toxic endogenous bacterial metabolites [45] which constitute potential PAMPs.

We documented the trend that high predicted BSL of the GM of allo-HSCT donors is associated with aGvHD, but more donor GM sequencing is required to demonstrate a significant correlation. Such a correlation would suggest donor antigen presenting cells or T-cells exposed to PAMPs prior to transplantation play a negative role post stemm cell infusion and that donor screening through 16S sequencing could be effective in selecting donors with low PAMP exposure.

### immuno-modulating metabolites

We predicted two metabolites known to influence intestinal barrier function, the AhR ligand tryptamine and the polyamine putrescine to be associated with non-aGvHD controls. In addition, we predicted a major propionate pathway to be decreased in aGvHD. Given the important difference in the GM composition between aGvHD cases and controls, many more predicted functional differences can be documented (results not shown), but these are at present relatively meaningless in the context of GvHD or allo-HSCT survival.

### T cells in aGvHD

Type 1 or Tc1/Th1 maturation is recognized as the dominant pattern in aGvHD and is linked to severe GI tract pathology. Th2 differentiation causes predominantly pathology in pulmonary, hepatic, and cutaneous tissues [46]. Also, Treg/Th17 decrease has been associated with aGvHD and proposed as a biomarker [22, 47]. Regulatory T-cell therapy for GvHD has been suggested [48, 49].

### Mortality associated species

The H2S producer and oral commensal *Atopobium parvulum* is the top mortality associated species. It has been found involved in oral infections [50]. Importantly, it is a predominant member of the adherent microbiome of pediatric IBD patients, associated with disease severity and promotes colitis in Il10-/- mice whilst eliciting a IL-17 response [51]. The probiotic bacterium *Bifidobacterium bifidum* induces Th17 differentiation [52]. *Clostridium aldenense* has been associated with a cytokine IL-7 response [53], i.e. generic development and maintenance of T cells. Likewise, *Phascolarctobacterium faecium* has been found to correlate positively with immune response, i.e. IFNγ^+^CD8^+^ T cells in a mouse model [54]. *Acidaminococcus fermentans* has been studied for its anaerobic glumate/glutamine fermentation. Glutamine has been shown to induce Treg cells in a mouse aGvHD model [55] and hence its catabolism would work against the induction of a Treg response.

All in all, the mortality associated species are associated with immune activation and an increased Th17/Treg ratio. In addition, *Lactobacillus gasseri* is a hydrogen peroxide producer. Although effective as an antimicrobial metabolite to control pathogens, hydrogen peroxide also functions as a damage associated molecular pattern (DAMP) in wound healing [56], which in the allo-HSCT setup could outweigh the benefits.

### aGvHD case and control associated species

*Staphylococcus epidermidis* is the most frequently isolated species of the coagulase negative *Staphylococci* from human stool and can cause infection in the gut [57], including necrotizing colitis in neonates [58]. Its position as the top aGvHD associated species in our findings is unexpected, but not illogical in a context of gut decontamination, given its presence in the hospital environment and its multiple antibiotic resistance [59]. *Raoultella ornithinolytica* is an emerging human pathogen with betalactamase based antibiotic resistance [60]. It is closely related to the pathogen *Klebsiella pneumoniae* which we also found associated with aGvHD. *Bacteroides ovatus* has been described as the predominant commensal intestinal microbe causing a systemic antibody response in inflammatory bowel disease [61]. *Rothia mucilaginosa* is a oral species associated with bacteremia in patients with neutropenia and leukemia [62]. Our data suggest translocation could occur in the gut. *Prevotella* species are associated with a Th17 response [63], with a pro-inflammatory role for the aGvHD associated species *Prevotella copri* in rheumatoid arthritis [64]. *Citrobacter koseri* is an opportunistic nosocomial pathogen, with high propensity to initiate brain abscesses during neonatal meningitis [65]. Like *Lactobacillus gasseri*, *Lactobacillus crispatus* is a producer of hydrogen peroxide, a potential DAMP signal. The position of the commensal *Bacteroides thetaiotaomicron* as an aGvHD associated species is intriguing. Although commensal, its mucolytic activity makes polysaccharides available for other organisms, including pathogens [66]. The top non-GvHD control associated *Bacteroides uniformis* induces IL-10 and Treg in PBMCs and LPMCs [67]. The position of *Barnesiella intestihominis* on the side of non-aGvHD controls is unexpected, since the bacterium has been shown to increase Th1 and decrease Treg response in cyclophosphamide anti-tumor therapy [68]. *Fusicatenibacter saccharivorans* has been reported decreased in active ulcerative colitis and induce IL-10 production by lamina propria mononuclear cells from not only colitis model mice but also UC patients [69]. *Flavonifactor plautii* has been shown to induce Treg cells in mice [70] and, importantly, the white blood cell growth factor GM-CSF in humans [53]. Lastly, we note the presence of the commensal butyrate producer *Anaerostipes caccae* among the non-aGvHD associated species.

The association of species with aGvHD cases and controls can be summarized as a pathogenic antibody eliciting set up on the side of the aGvHD cases and a commensal Treg cell eliciting set up on the control side.

### Modulating the allo-HSCT GM

From in silico allo-HSCT GM screening results, *Bifidobacterium longum* seems to present potential to modulate the allo-HSCT GM. Furthermore, *B. longum* elicits in majority IL-10 [52]. To increase putative case associated species exclusion, *B. longum* could be combined with another probiotic species, *Bifidobacterium breve*, with which it is found associated in vivo more frequently than respective prevalences predict, suggesting a synergistic relation. *B. breve* induces IL-10-producing Tr1 cells in the colon [71]. *Bacillus clausii*, which we find in a position similar to *B. longum* from a co-exclusion perspective, also presents interest in that it has been reported to increase IL-10 in allergic children [72]. All three species are predicted to respect the non-aGvHD control associated species we documented.

A selection of a high folate producers for *B. longum* and *B. breve* could be privileged for increased Treg stimulus [73]. Combination of *B. longum* and *B. breve* with a bifidogenic prebiotic seems indicated, either choosing or avoiding the butyrigenic inulin-type fructans (ITF) or arabinoxylan-oligosaccharides (AXOS) [74], depending on the patient’s aGvHD status [42, 43]. Other than folate (vitamin B9), supplementation with vitamin D to compensate deficiency in allo-HSCT [75, 76] is worth considering. In vitro metronidazole resistance among *Bifidobacteria* has been reported [77, 78]. *Bifidobacteria* are only moderately susceptible to the anti-gram positive vancomycin [77, 79, 80] and to the anti-anaerobe ceftazidime but susceptible to amoxicillin [77]. Of note, a rifaximin resistant strain of *B. longum* W11 has been documented and is commercially available [81]. Prophylactic use of rifaximin has been shown to lower IL-6 levels [82], preserve urinary 3-IS levels and increase survival in allo-HSCT as compared to ciprofloxacin/metronidazole [83]. *B. clausii* has a wide spectrum of natural antibiotic resistance, including macrolides, betalactams, aminoglycosides metronidazole, cefuroxime, ceftriaxone, cefotaxime and cephalosporin [84].

## Conclusions

Combination of several published and unpublished data sets and use of state-of-the-art bioinformatics algorithms has allowed us to document fine grain GM composition in relation to mortality and aGvHD. This in turn allowed for the assessment of the immune status of the GM based on documented immune profiles in literature of the species involved. Using in silico prediction of exclusion of mortality and aGvHD associated species, we could formulate a probiotic composition potentially competing with these species, whilst respecting non-aGvHD control associated species.

## Summary

- Immunosuppressive conditioning strength was found correlated with altered gut GM composition.
- The use of antibiotics during allo-HSCT can select resistant pathogens, which we found increased with aGvHD.
- Auto-FMT compensates bacterial diversity loss and lowers relative abundance of resistant pathogens.
- More donor stool sequencing is required to investigate the relation between aGvHD and donor exposure to pathobionts.
- Mortality and aGvHD associated species are predicted to elicit Th17 response whereas non-aGvHD control associated species are predicted to elicit a Treg response.
- *B. longum* and *B. breve* are predicted to compete with mortality and aGvHD associated species whilst respecting non-aGvHD control associated species.
- Antibiotics use can be tuned to correspond with Bifidobacterial resistance such that prophylactic supplementation during the antibiotics course becomes possible.
- Prebiotics and vitamins B9 and D can further enhance the prophylactic use of a supplement based on *B. longum* and *B. breve*.

## Data Availability

Data and ananlysis scripts have been made available under Github.

https://github.com/GeneCreek/GvHD-manuscript

## Declarations

## Acknowledgements

The authors acknowledge the contributions to the Short Read Archive made by the respective institutions and acknowledge scientific journals for enforcing this practice.

## Financial & competing interest disclosure

The authors are co-founders of and own shares in GeneCreek, Inc. Other than that, the authors declare no relevant affiliations or financial involvement with any organization or entity with a financial interest in or financial conflict with the subject matter or materials discussed in the manuscript. This includes employment, consultancies, honoraria, expert testimony, grants or patents received or pending, or royalties.

No writing assistance was utilized in the production of this manuscript.

## Ethical conduct of research

The authors declare that no human or animal experimental investigations have been carried out.

